# Effects of Glucagon-like Peptide-1 Receptor Agonists on Cardiovascular and Renal Outcomes: A Meta-Analysis and Meta-Regression Analysis

**DOI:** 10.1101/2021.10.14.21264803

**Authors:** Satoshi Yoshiji, Hiroto Minamino, Daisuke Tanaka, Shunsuke Yamane, Norio Harada, Nobuya Inagaki

## Abstract

**Aims:** Cardiovascular and renal effects of glucagon-like peptide-1 receptor agonists (GLP-1RAs) have been inconsistent in cardiovascular outcome trials, and factors associated with the efficacy of GLP-1RAs remain to be clarified. Here, we evaluate the cardiovascular and renal outcomes with GLP-1RAs and associations between these outcomes and HbA1c or weight reduction.

**Materials and Methods:** We searched PubMed/MEDLINE, EMBASE, and CENTRAL for randomized, placebo-controlled trials of GLP1-RAs reporting major adverse cardiovascular events (MACE; a composite of cardiovascular mortality, stroke, and myocardial infarction) as the primary outcome. We conducted a meta-regression analysis of primary and secondary outcomes with HbA1c or weight reduction following a meta-analysis with a random-effects model for these outcomes.

**Results:** We extracted data of 60,800 individuals from eight eligible studies (ELIXA, LEADER, SUSTAIN-6, EXSCEL, HARMONY, PIONEER 6, REWIND, and AMPLITUDE-O). GLP-1RAs reduced MACE (hazard ratio [HR] 0.86; 95% CI: 0.80–0.93; *P* < 0.001) and secondary outcomes including the composite renal outcome (0.80; 0.73–0.87; *P* < 0.001). In meta-regression analysis, every 1% reduction in HbA1c was associated with 26% and 35% decreases in the logarithm of HR of MACE (*P* = 0.044; *R*^*2*^ = 0.65) and the composite renal outcome (*P* = 0.040; *R*^*2*^ = 0.85), respectively. On the contrary, weight reduction was not associated with any outcome, including MACE (*P* = 0.390).

**Conclusions:** The reduction in HbA1c, but not body weight, is associated with cardiovascular and renal outcomes. The magnitude of HbA1c reduction can be a surrogate for cardiovascular and renal benefits of treatment with GLP-1RAs.

## Introduction

Major adverse cardiovascular events (MACE) and chronic kidney diseases are leading causes of morbidity and mortality in individuals with type 2 diabetes. Glucagon-like peptide-1 receptor agonists (GLP-1RAs) have shown cardiovascular and renal benefits in cardiovascular outcome trials and played a key role in the management of type 2 diabetes.^1, 2^

To date, a series of randomized, placebo-controlled clinical trials on cardiovascular and renal outcomes with GLP-1RAs has been reported, including ELIXA with lixisenatide, LEADER with liraglutide, SUSTAIN-6 with semaglutide, EXSCEL with exenatide once weekly, HARMONY with albiglutide, PIONEER 6 with oral semaglutide, REWIND with dulaglutide, and AMPLITUDE-O with efpeglenatide.^3–10^ These trials evaluated the safety and efficacy of GLP-1RAs regarding the primary composite MACE outcome (cardiovascular mortality, stroke, or myocardial infarction), components of MACE, hospitalization due to heart failure, all-cause mortality, and renal outcomes. GLP-1RAs consistently showed cardiovascular and renal safety, but the cardiovascular and renal efficacies were inconsistent across the trials. Thus, further studies are warranted to elucidate factors associated with cardiovascular and renal benefits of GLP-1RAs.

Regarding the mechanism of action for cardiovascular and renal efficacies, it has been reported that reducing HbA1c or body weight contributes to cardiovascular, renal, and mortality risk reduction in legacy studies.^11–16^ However, although cardiovascular outcome trials of GLP-1RAs have reported an effective reduction in HbA1c and body weight, it remains unclear whether HbA1c or body weight reduction is associated with the cardioprotective and renoprotective profiles.

To examine the overall cardiovascular and renal efficacies of GLP-1RAs, we performed a meta-analysis of cardiovascular outcome trials of GLP-1RAs. We then conducted a meta-regression analysis to evaluate whether the reduction in HbA1c or body weight is associated with cardiovascular and renal efficacies of GLP-1RAs.

## Methods

### Data Sources and Search

We performed a data search in accordance with the Preferred Reporting Items for Systematic Reviews and Meta-Analysis (PRISMA) statement.^17^ We searched the PubMed/MEDLINE, EMBASE, and CENTRAL databases for eligible randomized, placebo-controlled trials of GLP-1RAs reporting MACE in individuals with type 2 diabetes, published until January 23, 2022. We placed no restrictions on the study population or language. The following search terms were used: “glucagon-like peptide-1 receptor agonist,” “GLP-1 receptor agonist,” “lixisenatide,” “liraglutide,” “semaglutide,” “exenatide,” “albiglutide,” “liraglutide,” “oral semaglutide,” “efpeglenatide,” “type 2 diabetes,” “randomized controlled trial,” “randomized placebo-controlled trial,” “Major adverse cardiovascular events,” “MACE,” “cardiovascular mortality,” “myocardial infarction,” “acute coronary syndrome,” “cardiovascular,” “stroke,” “death,” “heart failure,” “kidney disease,” and “renal.” The PRISMA flow diagram is presented in **Figure 1**. The protocol for this study is registered in PROSPERO (CRD42021273058).

**Figure 1.**
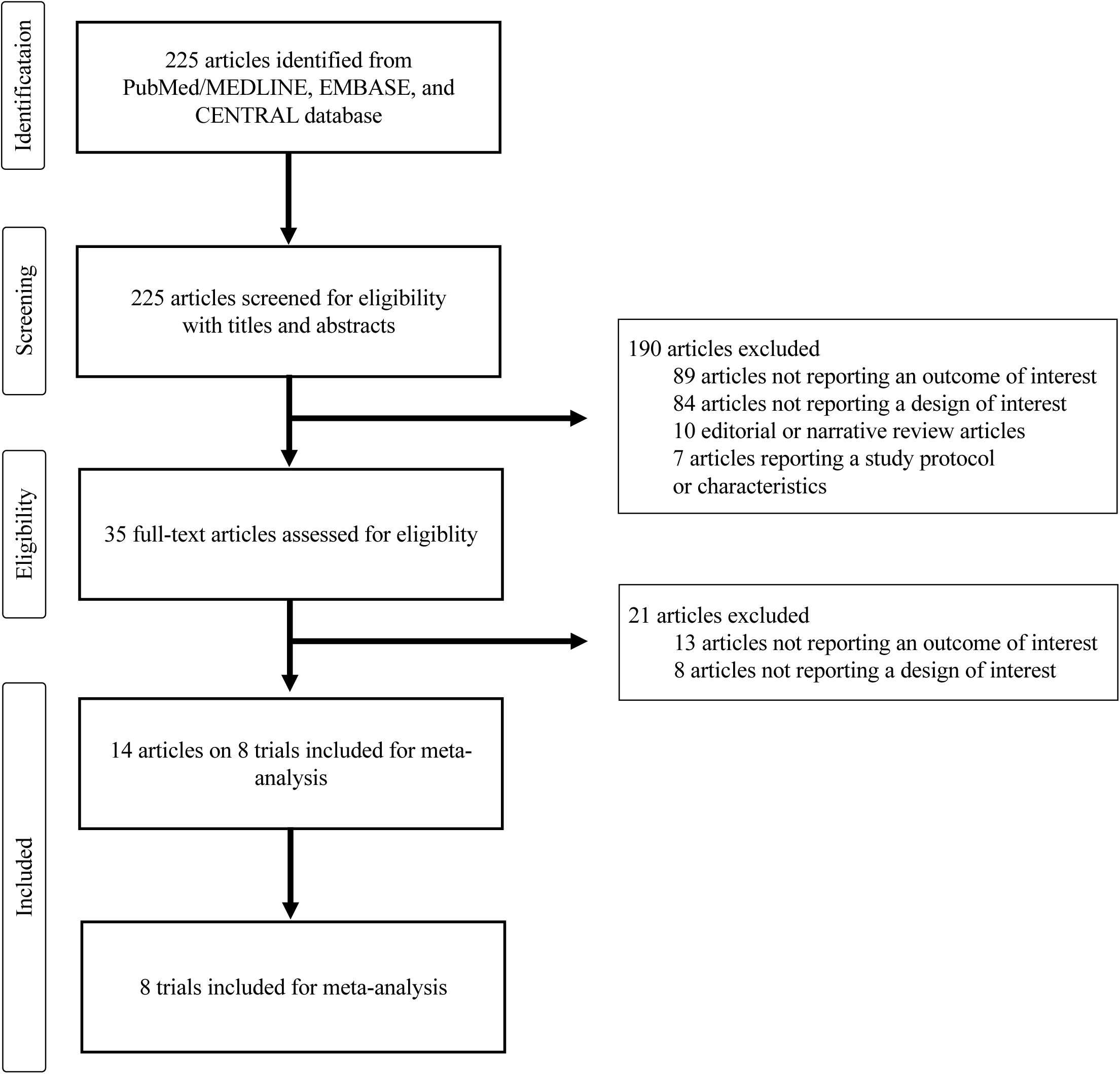
PRISMA flow diagram for identification of studies.

### Study Selection

We selected studies reporting MACE as the primary outcome.

### Data Extraction and Quality Assessment

We extracted summary statistics of the eligible studies from published articles of the trials and their supplementary materials. We did not seek individual-level data. For the primary and secondary outcomes, we obtained relevant hazard ratio (HR) and 95% confidence interval (95% CI) data. We evaluated the risk of bias for eligible studies with the Cochrane Risk of Bias Tool.^18^ SY and HM conducted data extraction and quality assessment. SY and HM independently conducted the search and solved any discordance through discussion.

### Data Synthesis and Analysis

The primary outcome was the HR of MACE. The secondary outcomes were the HR of individual components of MACE, all-cause mortality, hospitalization due to heart failure, composite renal outcome, and renal function outcome. The definitions of “composite renal outcome” and “renal function outcome” were not standardized across the trials; detailed definitions of these renal outcomes in each trial are provided in **Supplementary Table 1**.

For the meta-analysis, we used the inverse variance method with a random-effects model to obtain a weighted average of HR with 95% CI for each outcome. We used the DerSimonian-Laird estimator for the assessment of between-study variance.^19^ We used the *I*^*2*^ index and Cochran’s Q test to measure the degree of heterogeneity. The degree of heterogeneity based on the *I*^*2*^ index was defined as low (*I*^*2*^ index ≤ 25%), moderate (*I*^*2*^ index 26–50%), and high (*I*^*2*^ index > 50%).^20^ Results with *P* < 0.05 in Cochran’s Q test represent significant heterogeneity between the studies.

Regarding meta-regression analysis, normalized HbA1c and body weight reduction by each GLP-1RA were calculated from head-to-head clinical trials as described previously.^21^ For normalization, we defined HbA1c reduction and body weight reduction of liraglutide as 1.0% and 1.0 kg, respectively, considering that liraglutide was compared head-to-head against all the other GLP-1RAs.^22–28^ Using the following equations (1) and (2), we normalized the HbA1c and body weight reduction of each GLP-1RA against those of liraglutide by taking the ratio of their %HbA1c reduction and %body weight reduction from baseline, respectively (**Supplementary Table 3**):

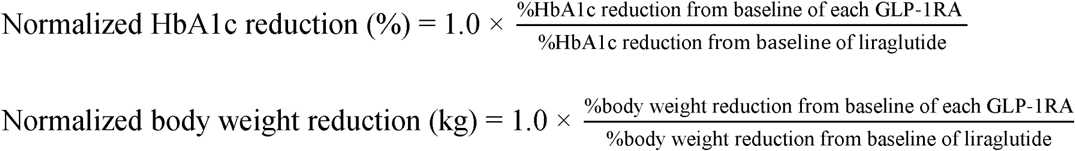

Using either normalized HbA1c change or body weight change as an independent variable, we conducted univariable meta-regression analyses with a mixed-effects model for cardiovascular and renal outcomes with the logarithm of HR (log-HR) as a dependent variable. Results with *P* < 0.05 were considered significant.

We performed all statistical analyses with R (version 4.0.2), using the R packages meta (version 4.18.2) and metafor (version 3.02).

## Results

### Study identification

Among the 225 articles screened with the prespecified screening strategy, eight randomized, placebo-controlled cardiovascular outcome trials reporting MACE as the primary outcome were identified. Data from the following eight trials involving 60,080 individuals were analyzed in the present meta-analysis: ELIXA with lixisenatide, LEADER with liraglutide, SUSTAIN-6 with semaglutide, EXSCEL with exenatide once weekly, HARMONY with albiglutide, PIONEER 6 with oral semaglutide, REWIND with dulaglutide, and AMPLITUDE-O with efpeglenatide. The study selection process based on PRISMA is presented in **Figure 1**. According to the Cochrane risk of bias tool,^18^ the risk of bias for all included trials was evaluated as low (**Supplementary Table 2**).

For all trials except for ELIXA, the primary outcome was MACE, which was the time to the first occurrence of cardiovascular mortality, stroke, or myocardial infarction. For ELIXA, the primary outcome was extended MACE, which was MACE with an additional component, i.e., hospitalization due to unstable angina. However, hospitalization due to unstable angina accounted for less than 2.5% of the primary outcome in ELIXA. Considering that hospitalization for unstable angina had a minimal influence on the primary outcome, we included ELIXA in the meta-analysis and meta-regression analysis. HARMONY and PIONEER 6 did not report renal outcomes and were excluded from the analysis of renal outcomes.

### Baseline characteristics

The baseline characteristics and other details of the eight trials are summarized in **Table 1**. The median duration of follow-up was the shortest in PIONEER 6 (1.3 years) and the longest in REWIND (5.4 years). The average age of patients ranged from 59.9 years in ELIXA to 66.2 years in REWIND. Of the 60,080 individuals, 21,930 (37%) were women, and 45,640 (76%) were of white ethnicity. The average body mass index varied from 30.1 kg/m^2^ (ELIXA) to 32.8 kg/m^2^ (SUSTAIN-6), average baseline HbA1c ranged from 7.3% (REWIND) to 8.9% (AMPLITUDE-O), and average estimated glomerular filtration rate (eGFR) from 74 mL/min per 1.73 m^2^ (PIONEER 6) to 80 mL/min per 1.73 m^2^ (LEADER and SUSTAIN-6). The proportion of individuals with previously established cardiovascular disease was 77% (46,105 out of 60,080 individuals) across the eight trials and ranged from 31% (REWIND) to 100% (ELIXA and HARMONY).

**Table 1.**
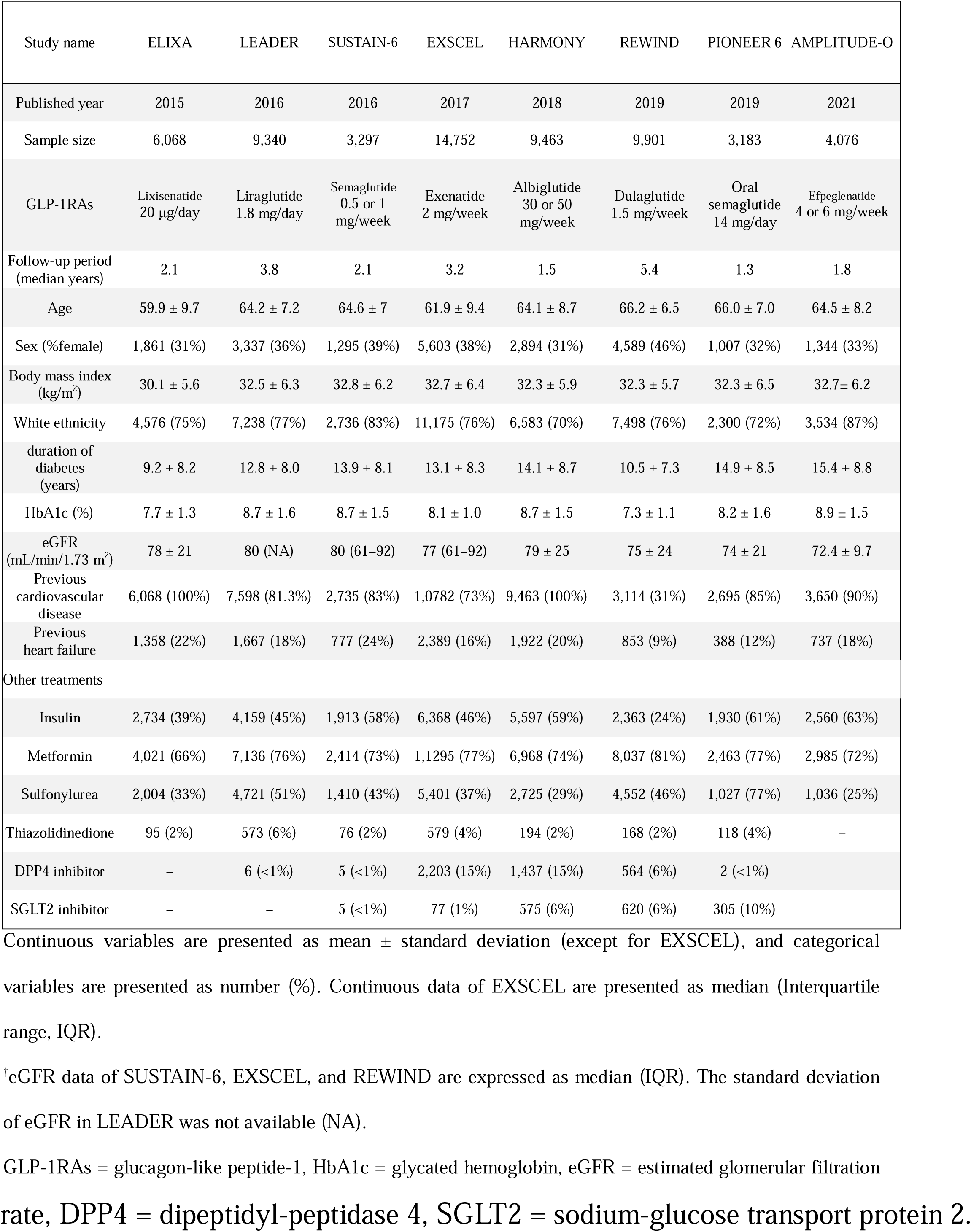
Study characteristics.

### Meta-analysis

Overall, GLP-1RAs reduced MACE (HR: 0.86; 95% CI: 0.80–0.93; *P* < 0.001; *I*^*2*^ = 44.5%) (**Figure 2**). As for the individual components of MACE, GLP-1RAs reduced cardiovascular mortality (HR: 0.87; 95% CI: 0.80–0.94; *P* = 0.001; *I*^*2*^ = 12.8%), myocardial infarction (HR: 0.90; 95% CI: 0.83–0.98; *P* = 0.020; *I*^*2*^ = 27.4%), and stroke (HR: 0.83; 95% CI: 0.76–0.92; *P* < 0.001; *I*^*2*^ = 0%).

**Figure 2.**
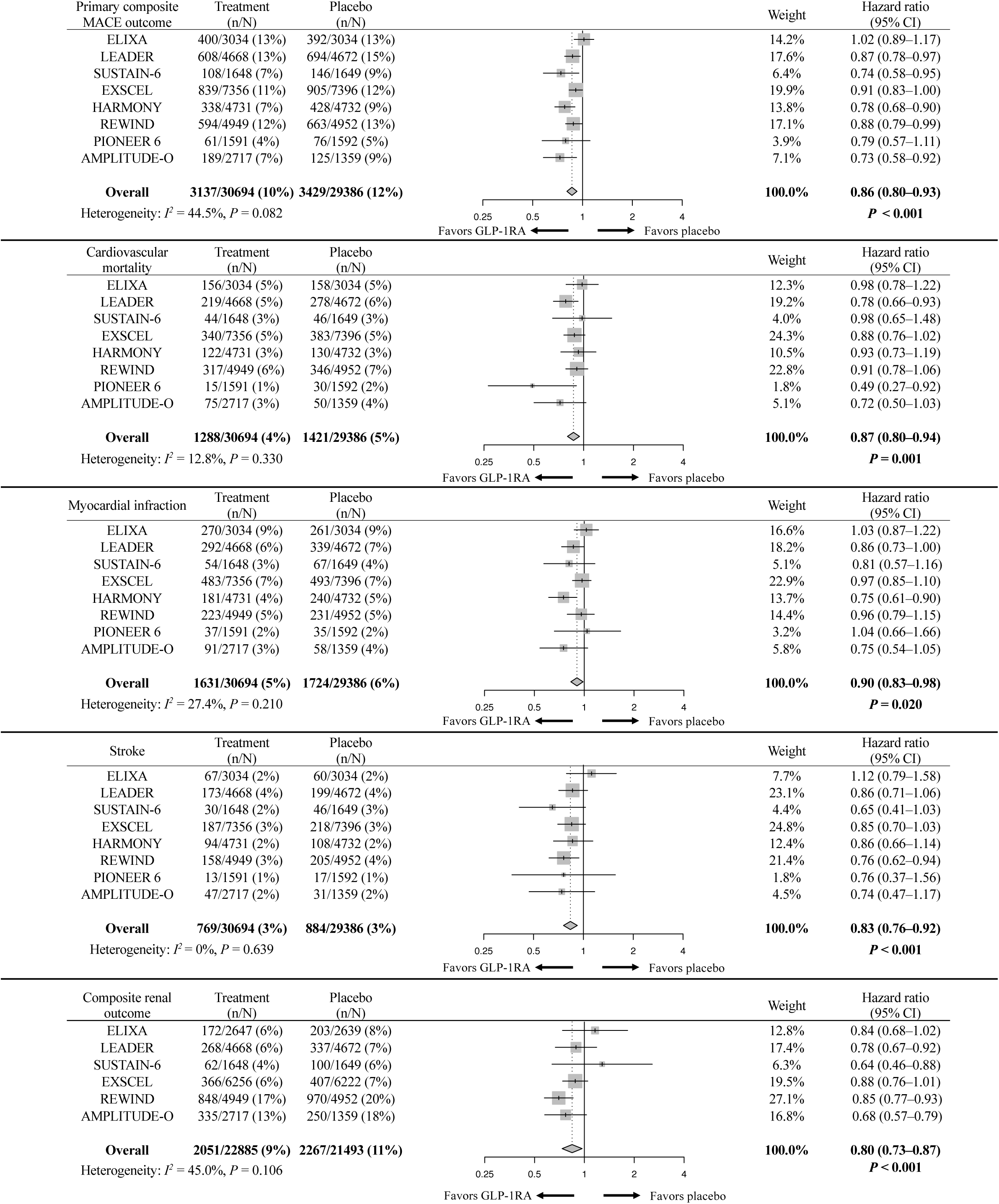
Effects of GLP-1RAs on three-point MACE outcome, cardiovascular mortality, myocardial infarction, stroke, and composite renal outcome. MACE = major adverse cardiovascular events.

As for the other secondary outcomes, GLP-1RAs reduced composite renal outcome (HR: 0.80; 95% CI: 0.73–0.87; *P* < 0.001; *I*^*2*^ = 45.0%) (**Figure 2**), all-cause mortality (HR: 0.88; 95% CI: 0.82–0.94; *P* < 0.001; *I*^2^ = 10.5%), hospitalization due to heart failure (HR: 0.89; 95% CI: 0.82–0.98; *P* = 0.013; *I*^*2*^ = 2.5%), and renal function outcome (HR: 0.84; 95% CI: 0.73–0.97; *P* = 0.016; *I*^*2*^ = 31.4%) (**Supplementary Figure 1**).

### Subgroup analysis

In a subgroup analysis of MACE stratified by the structural backbones of GLP-1RAs (**Supplementary Figure 2**), exendin-based GLP-1RAs did not reduce MACE with high heterogeneity (HR: 0.90; 95% CI: 0.78–1.04; *P* = 0.163; *I*^*2*^ = 67.2%). On the contrary, human GLP-1-based GLP-1RAs reduced MACE with low heterogeneity (HR: 0.84; 95% CI: 0.79–0.90; *P* < 0.001).

In another subgroup analysis of MACE stratified by the history of established cardiovascular disease (**Supplementary Figure 3**), GLP-1RAs reduced MACE in individuals with established cardiovascular disease (HR: 0.85; 95% CI: 0.79–0.92; *P* < 0.001; *I*^*2*^ = 48.0%) but not in those without established cardiovascular disease (HR: 0.94; 95% CI: 0.83–1.06; *P* = 0.303; *I*^*2*^ = 0.0%).

### Meta-regression analysis

Normalized HbA1c and body weight reduction used in the meta-regression analysis are presented in **Supplementary Table 3**. The meta-regression analysis of the eight pooled trials showed that normalized HbA1c reduction was associated with the log-HR of MACE (slope: – 0.26; 95% CI: –0.52 to –0.01; *P* = 0.044; *R*^*2*^ = 0.65); i.e., every 1% reduction in HbA1c was associated with a 26% decrease in log-HR of MACE (*P* = 0.044). On the contrary, normalized body weight reduction showed no association with the log-HR of MACE (slope: –0.05; 95% CI: –0.17 to 0.07; *P* = 0.390; *R*^*2*^ < 0.01) (**Figure 3**). Among the secondary outcomes, normalized HbA1c reduction was associated with the log-HR of the composite renal outcome (slope: –0.35; 95% CI: –0.68 to –0.02; *P* = 0.040; *R*^*2*^ = 0.85) (**Figure 4**), which means that every 1% reduction in HbA1c was associated with 35% decrease in the logarithm of HR (log-HR) of MACE. Although the associations between normalized HbA1c reduction and other secondary outcomes were not significant, all outcomes except hospitalization due to heart failure showed negative slopes for the regression line, which was directionally concordant with MACE. As for body weight, normalized body weight reduction was not associated with any secondary outcome; the 95% CIs were wide and ranged from negative to positive values (**Figure 4** and **Supplementary Figure 4**).

**Figure 3.**
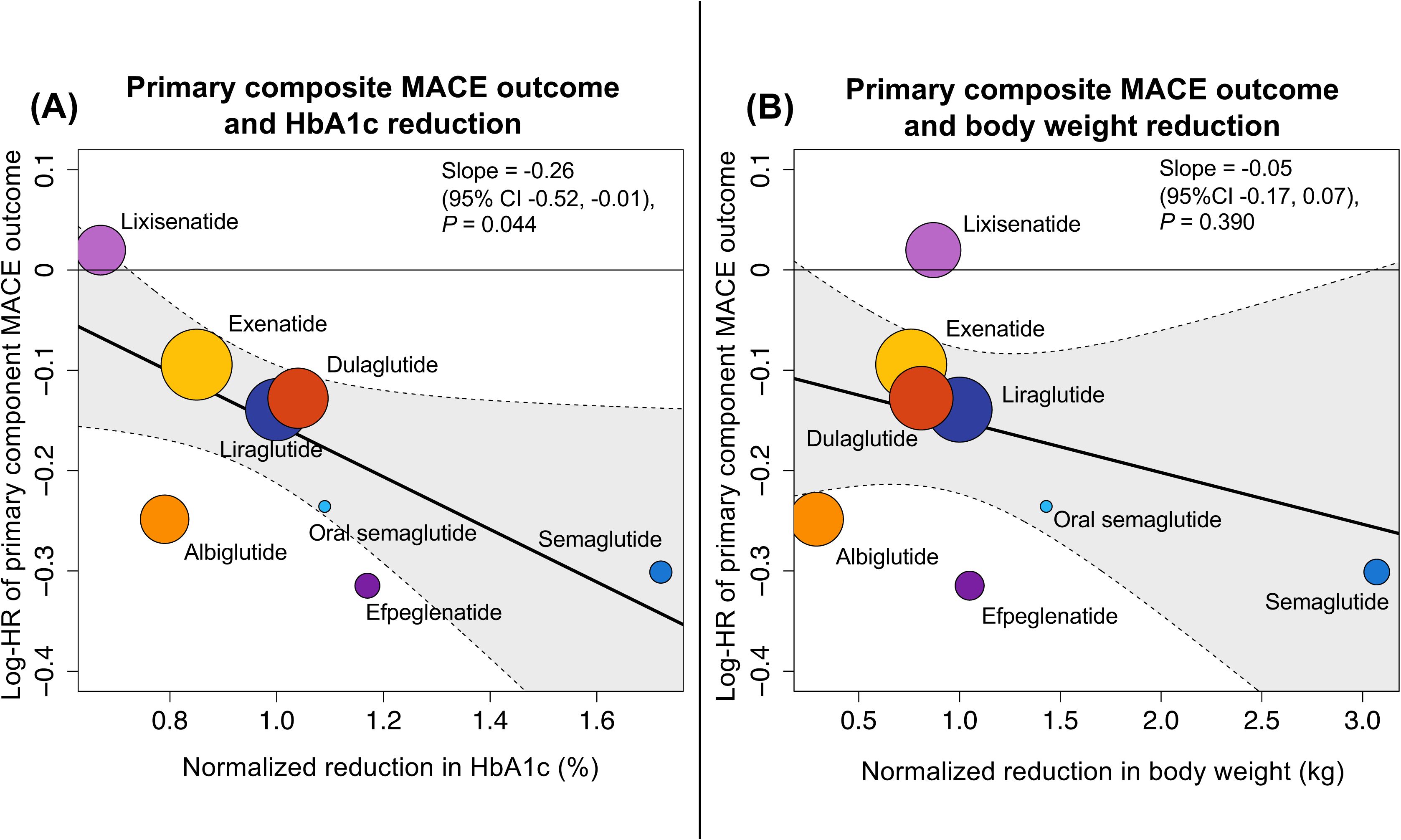
Univariable meta-regression analysis of normalized reduction in HbA1c (A) or body weight (B) with the logarithm of hazard ratio (log-HR) for the primary composite MACE outcome. The size of each trial’s circle is inversely proportional to the variance of hazard ratio. MACE = major adverse cardiovascular events.

**Figure 4.**
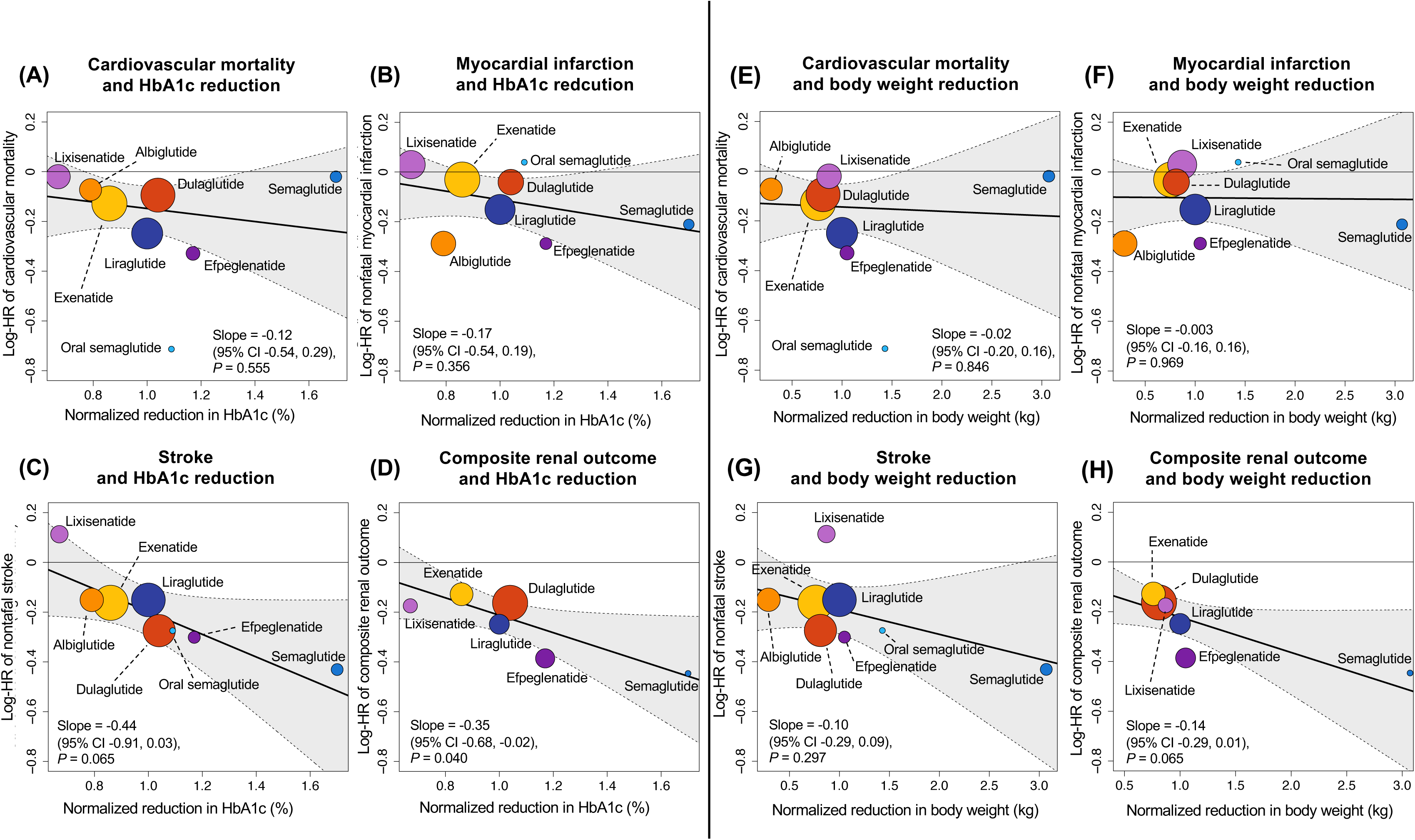
Univariable meta-regression analysis of normalized reduction in HbA1c reduction (A– D) or body weight (E–H) with the logarithm of hazard ratio (log-HR) for cardiovascular mortality (A, E), myocardial infarction (B, F), stroke (C, G), and composite renal outcome (D, H). The size of each trial’s circle is inversely proportional to the variance of hazard ratio.

## Discussion

The present meta-analysis and meta-regression analysis involved 60,080 individuals from eight cardiovascular outcome trials, including the latest evidence from AMPLITUDE-O. In the meta-analysis, GLP-1RAs reduced MACE, its components, cardiovascular mortality, myocardial infarction, stroke, all-cause mortality, hospitalization due to heart failure, the composite renal outcome, and the renal function outcome. In meta-regression analysis, a reduction in HbA1c was associated with a decrease in log-HR of MACE and the composite renal outcome. On the contrary, weight reduction was not associated with any outcome. To the best of our knowledge, this is the largest study combining meta-analysis and meta-regression analysis to evaluate cardiovascular and renal efficacies of GLP-1RAs.

Our meta-analysis reaffirms the cardiovascular and renal benefits of GLP-1RAs suggested by previous studies^29–31^ and supports the current guidelines that recommend GLP-1RAs for the treatment of diabetes in individuals with established cardiovascular diseases or in those who are at high risk of cardiovascular diseases.^1, 2, 32, 33^ In addition to the trials included in previous meta-analyses, our study also included AMPLITUDE-O, which is the latest trial using the GLP-1RA with the exendin backbone to show the successful reduction of MACE. By including AMPLITUDE-O, our meta-analysis increased the sample size by 4,076, providing more robust evidence on GLP-1RA efficacy.

In a subgroup analysis stratified by the history of established cardiovascular disease (Supplementary Figure 2), GLP-1RAs reduced MACE in individuals with established cardiovascular disease but not in those without established cardiovascular disease. Taking into account the anti-atherosclerotic properties of GLP-1RAs,^34^ it is conceivable that the cardiovascular benefits of GLP-1RAs were more prominent in individuals with established cardiovascular disease.

In terms of individual study details, all studies, except for ELIXA and EXSCEL, reported a reduction in MACE. Notably, in AMPLITUDE-O, efpeglenatide reduced MACE for the first time as an exendin-based GLP-1RA. Based on the findings from AMPLITUDE-O, we conducted the subgroup analysis of MACE according to the structural backbone of GLP-1RAs (Supplementary Figure 3). While human GLP-1-based GLP-1RAs reduced MACE (HR: 0.84; 95% CI: 0.79–0.90; *P* < 0.001) with low heterogeneity (*I*^*2*^ = 0.0%, *P* = 0.513), exendin-based GLP-1RAs did not reduce MACE (HR: 0.90; 95% CI: 0.78–1.04; *P* = 0.163). The difference could be due to the large heterogeneity of exendin-based GLP-1 trials (*I*^2^ = 67.2%, *P* = 0.047). Individually, ELIXA was different from the other two trials (EXSCEL and AMPLITUDE-O) in terms of recruited individuals with a recent acute coronary syndrome.^3^ It should also be noted that EXSCEL reported poor adherence to the drug (around 40% of premature discontinuation of the trial) and that lixisenatide, used in ELIXA, has a half-life of 2–4 h, which is considerably shorter than that of other GLP-1RAs.^35^ Therefore, in ELIXA and EXSCEL, multiple factors such as study design, adherence to the drug, and half-life could have influenced the cardiovascular outcomes. On the contrary, in AMPLITUDE-O, a reduction in MACE and other secondary outcomes was achieved despite the shortest follow-up period of 1.8 years (2.1 years for ELIXA and 3.2 years for EXSCEL),^3, 6, 10^ showing that exendin-based GLP-1RAs can also exert cardioprotective and renoprotective effects.

Regarding the underlying mechanism of GLP-1RA efficacy, factors associated with the cardioprotective and renoprotective profiles of GLP-1RAs remained to be clarified.^36^ Therefore, to determine whether the reduction in HbA1c or body weight (two key clinical variables decreased by GLP-1RAs) is associated with the cardioprotective and renoprotective effects of GLP-1RAs, we conducted a meta-regression analysis of the log-HR of MACE and other secondary outcomes with normalized reduction in HbA1c or body weight.

As for the primary composite MACE outcome (**Figure 3**), the results showed that the log-HR of MACE decreased by 26% for every 1.0% normalized HbA1c reduction. In contrast, log-HR of MACE was not associated with normalized body weight reduction. In individual studies, albiglutide caused minimal weight reduction but still showed a reduction in log-HR of MACE. Contrarily, lixisenatide, which showed a greater normalized body weight reduction than albiglutide and dulaglutide, did not reduce cardiovascular risk.

The present finding that only HbA1c reduction, but not body weight reduction, is associated with cardiovascular risk is consistent with that of a recent post-hoc mediation study of the LEADER trial.^37^ The mediation analysis showed that HbA1c was potentially the largest mediator of cardiovascular efficacy of liraglutide, with percentage mediation being 41–83%, whereas body weight was calculated to have a considerably lower percentage mediation of 4– 14%. Considering the findings of our meta-regression analysis and the mediation analysis, it seems likely that HbA1c reduction can be a marker of cardiovascular efficacy of GLP-1RAs.

However, whether HbA1c reduction constitutes a direct contributor or represents unmeasured contributors remains to be clarified. In addition to glucose-lowering effects, pleiotropic effects, including amelioration of endothelial damage and chronic inflammation,^38, 39^ are potential contributors to cardiovascular risk reduction with GLP-1RAs.

As for the secondary outcomes, every 1.0% normalized HbA1c reduction was associated with a 35% decrease in log-HR of the composite renal outcome. On the contrary, normalized body weight reduction presented a high variability among studies and was not associated with any secondary outcome (**Figure 4** and **Supplementary Figure 2**). Consistent with our finding that the reduction in HbA1c, but not body weight, is associated with the composite renal outcome, another mediation analysis of the LEADER and SUSTAIN 6 trials on the composite renal outcome suggested that HbA1c is potentially the greatest mediator of renal benefits, whereas body weight has little to no effect.^39^ In contrast to the association of HbA1c reduction with the renal composite outcome, we did not find a significant association with the renal function outcome (the narrower renal endpoint). This discrepancy may be explained by the definitions of these outcomes: the renal composite outcome included new-onset macroalbuminuria, which is predominantly prevented by GLP-1RAs,^40^ whereas the renal function outcome did not (**Supplementary Table 1**). However, we must note that cardiovascular outcome trials were not designed to detect renal events as the primary outcome. In this regard, the ongoing FLOW trial (NCT03819153),^41^ which evaluates the effects of semaglutide on hard renal endpoints as the primary outcome in individuals with chronic kidney disease, is expected to provide more insights into the renal effects of the GLP-1RAs.

This study has some limitations. First, we used only summary data for meta-analysis and did not use individual-level data. Second, although heterogeneity between studies was generally low for most of the outcomes, moderate heterogeneity was observed for the renal composite outcome and the renal function. This is likely because definitions of renal outcomes were not standardized across the trials (**Supplementary Table 1**). Third, although we conducted the meta-analysis and meta-regression analyses on various secondary outcomes in addition to the primary outcome, the included trials were not sufficiently powered to detect the differences in the secondary outcomes. This may have reduced the statistical power for meta-regression analysis of the secondary outcomes.

In conclusion, GLP-1RAs reduce MACE, its components, all-cause mortality, hospitalization due to heart failure, the composite renal outcome, and the renal function outcome.

The meta-regression analysis showed that the reduction in HbA1c, but not body weight, is associated with cardiovascular and renal outcomes of GLP-1RAs. The magnitude of HbA1c reduction can be a surrogate for cardiovascular and renal benefits of treatment with GLP-1RAs.

## Data Availability

All the data were extracted from publicly available sources and included in the present article.

