## Supplementary material for "Effects of Glucagon-like Peptide-1 Receptor Agonists on Cardiovascular and Renal Outcomes: A Meta-Analysis and Meta-Regression Analysis"

Supplementary Table 1. Definition of renal outcomes.

Supplementary Table 2. Risk of bias assessment.

Supplementary Table 3. Normalized HbA1c and body weight reduction used in the meta-regression analysis.

Supplementary Figure 1. Meta-analysis of all-cause mortality, hospitalization due to heart failure, and renal function outcome.

Supplementary Figure 2. Subgroup analysis of effects of GLP-1RAs on the primary composite MACE outcome stratified by prior history of established cardiovascular disease.

Supplementary Figure 3. Subgroup analysis of effects of GLP-1RAs on the primary composite MACE outcome stratified by structural similarities.

Supplementary Figure 4. Univariable meta-regression analysis of normalized reduction in HbA1c or body weight with the logarithm of hazard ratio for all-cause mortality, hospitalization due to heart failure, and renal function outcome.

**Supplementary Table 1.** Definition of renal outcomes.

| Trial | Renal function outcome | Composite renal outcome |
|---|---|---|
| ELIXA | Doubling of serum creatinine | New-onset macroalbuminuria |
| LEADER | Doubling of serum creatinine | New-onset macroalbuminuria, doubling of serum creatinine (eGFR < 45 mL/min per $1.73m^2$), renal replacement therapy, death due to kidney disease |
| SUSTAIN-6 | Doubling of serum creatinine | New-onset macroalbuminuria, doubling of serum creatinine (eGFR < 45 mL/min per $1.73m^2$), renal replacement therapy, death due to kidney disease |
| EXSCEL | ≥ 40% worsening of eGFR, renal-replacement therapy, death due to kidney disease | New-onset persistent macroalbuminuria, ≥ 40% worsening of eGFR, renal replacement therapy, death due to kidney disease |
| HARMONY | Not reported | Not reported |
| REWIND | ≥ 40% worsening of eGFR | New-onset macroalbuminuria, ≥ 30% worsening of eGFR, renal replacement therapy |
| PIONEER 6 | Not reported | Not reported |
| AMPLITUDE-O | ≥ 40% worsening of eGFR for ≥ 30 days, renal replacement therapy for ≥ 90 days, or eGFR < 15 ml/min/1.73 $m^2$ for ≥ 30 days, or all-cause death | New-onset macroalbuminuria, increase in urinary albumin-to-creatinine ratio of ≥30 % from baseline, a sustained decrease in the eGFR of at least 40% for ≥ 30 days, renal-replacement therapy for ≥ 90 days, and a sustained eGFR of ≤ 15 ml/min/1.73 $m^2$ for ≥30 days |

eGFR = estimated glomerular filtration rate.

**Supplementary Table 2.** Risk of bias assessment.

| | ELIXA | LEADER | SUSTAIN-6 | EXSCEL | HARMONY | REWIND | PIONEER-6 | AMPLITUDE-O |
|---|---|---|---|---|---|---|---|---|
| Random sequence generation (selection bias) | Low risk | Low risk | Low risk | Low risk | Low risk | Low risk | Low risk | Low risk |
| Allocation concealment (selection bias) | Low risk | Low risk | Low risk | Low risk | Low risk | Low risk | Low risk | Low risk |
| Blinding of participants and researchers (performance bias) | Low risk | Low risk | Low risk | Low risk | Low risk | Low risk | Low risk | Low risk |
| Blinding of outcome assessment (detection bias) | Low risk | Low risk | Low risk | Low risk | Low risk | Low risk | Low risk | Low risk |
| Incomplete outcome data (attrition bias) | Low risk | Low risk | Low risk | Low risk | Low risk | Low risk | Low risk | Low risk |
| Selective reporting (reporting bias) | Low risk | Low risk | Low risk | Low risk | Low risk | Low risk | Low risk | Low risk |
| Other bias | Low risk | Low risk | Low risk | Low risk | Low risk | Low risk | Low risk | Low risk |

**Supplementary Table 3.** Normalization of HbA1c and body weight reduction using data from head-to-head trials.

| – | – | Drug | Exenatide once weekly | Dulaglutide | Albiglutide | Lixisenatide | Oral semaglutide | Efpeglenatide | Semaglutide |
|---|---|---|---|---|---|---|---|---|---|
| References for head-to-head trials | – | – | 22 | 23 | 24 | 25 | 26 | 27 | 28 |
| Reduction in head-to-head trials | %HbA1c reduction from baseline | GLP-1RA compared against liraglutide | 15.06 | 17.53 | 9.51 | 14.29 | 15.00 | 20.13 | 20.73 |
| | | Liraglutide | 17.62 | 16.79 | 12.07 | 21.43 | 13.75 | 17.25 | 12.05 |
| | %body weight reduction from baseline | GLP-1RA compared against liraglutide | 2.95 | 3.09 | 1.95 | 3.68 | 4.06 | 3.80 | 6.00 |
| | | Liraglutide | 3.88 | 3.82 | 6.68 | 4.22 | 2.84 | 3.61 | 1.95 |
| ↓Normalize reduction in HbA1c and body weight of each GLP-1RA against those of liraglutide, which were defined as 1.0% and 1.0 kg, respectively. | | | | | | | | | |
| Normalized reduction | Normalized HbA1c reduction | GLP-1RA normalized against liraglutide | 0.85 | 1.04 | 0.79 | 0.67 | 1.09 | 1.17 | 1.72 |
| | | Liraglutide | 1.00 | 1.00 | 1.00 | 1.00 | 1.00 | 1.00 | 1.00 |
| | Normalized body weight reduction | GLP-1RA normalized against liraglutide | 0.76 | 0.81 | 0.29 | 0.87 | 1.43 | 1.05 | 3.07 |
| | | Liraglutide | 1.00 | 1.00 | 1.00 | 1.00 | 1.00 | 1.00 | 1.00 |

**Formula used for normalizing HbA1c and body weight reduction**

$$\text{Normalized HbA1c reduction (\%)} = 1.0\ (\%) \times \frac{\text{\%HbA1c reduction from baseline of a each GLP−1RA in a head−to−head trial}}{\text{\%HbA1c reduction of liraglutide in a head−to−head trial}}$$

$$\text{Normalized body weight reduction (kg)} = 1.0\ (\text{kg}) \times \frac{\text{\%body weight reduction from baseline of a each GLP−1RAa head−to−head trial}}{\text{\%body weight reduction from baseline of liraglutide in a head−to−head trial}}$$

GLP-1RA = glucagon-like peptide-1 receptor agonist.

**Supplementary Figure 1.** Meta-analysis of all-cause mortality, hospitalization due to heart failure, and renal function outcome.

| All-cause mortality | Treatment (n/N) | Placebo (n/N) | Weight | Hazard ratio (95% CI) |
|---|---|---|---|---|
| ELIXA | 211/3034 (7%) | 223/3034 (7%) | 11.2% | 0.94 (0.78–1.13) |
| LEADER | 381/4668 (8%) | 447/4672 (10%) | 19.3% | 0.85 (0.74–0.97) |
| SUSTAIN-6 | 62/1648 (4%) | 60/1649 (4%) | 3.3% | 1.05 (0.74–1.50) |
| EXSCEL | 507/7356 (7%) | 584/7396 (8%) | 25.0% | 0.86 (0.77–0.97) |
| HARMONY | 196/4731 (4%) | 205/4732 (4%) | 10.5% | 0.95 (0.79–1.16) |
| REWIND | 536/4949 (11%) | 592/4952 (12%) | 25.0% | 0.90 (0.80–1.01) |
| PIONEER 6 | 23/1591 (1%) | 45/1592 (3%) | 1.7% | 0.51 (0.31–0.84) |
| AMPLITUDE-O | 111/2717 (4%) | 69/1359 (5%) | 4.5% | 0.78 (0.58–1.06) |
| **Overall** | **2027/30694 (7%)** | **2225/29386 (8%)** | **100.0%** | **0.88 (0.82–0.94)** **$P < 0.001$** |

Heterogeneity: $I^2 = 10.5\%$, $P = 0.349$

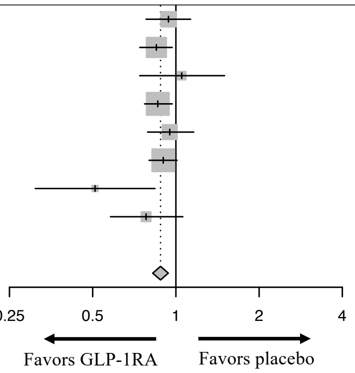


| Hospitalization due to heart failure | Treatment (n/N) | Placebo (n/N) | Weight | Hazard ratio (95% CI) |
|---|---|---|---|---|
| ELIXA | 122/3034 (4%) | 127/3034 (4%) | 12.6% | 0.96 (0.75–1.23) |
| LEADER | 218/4668 (5%) | 248/4672 (5%) | 22.8% | 0.87 (0.73–1.05) |
| SUSTAIN-6 | 59/1648 (4%) | 54/1649 (3%) | 5.8% | 1.11 (0.77–1.61) |
| EXSCEL | 219/7356 (3%) | 231/7396 (3%) | 22.0% | 0.94 (0.78–1.13) |
| HARMONY | 79/4731 (2%) | 111/4732 (2%) | 9.5% | 0.71 (0.53–0.94) |
| REWIND | 213/4949 (4%) | 226/4952 (5%) | 21.5% | 0.93 (0.77–1.12) |
| PIONEER 6 | 21/1591 (1%) | 24/1592 (2%) | 2.3% | 0.86 (0.48–1.55) |
| AMPLITUDE-O | 40/2717 (1.5%) | 31/1359 (2.3%) | 3.5% | 0.61 (0.38–0.98) |
| **Overall** | **971/30694 (3%)** | **1052/29386 (4%)** | **100.0%** | **0.89 (0.82–0.98)** **$P = 0.013$** |

Heterogeneity: $I^2 = 2.5\%$, $P = 0.410$

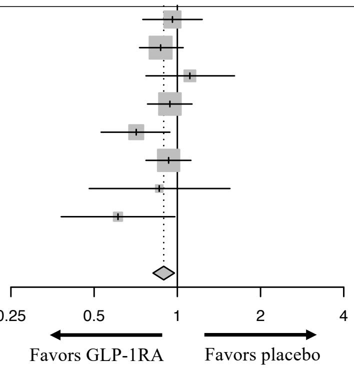


| Renal function outcome | Treatment (n/N) | Placebo (n/N) | Weight | Hazard ratio (95% CI) |
|---|---|---|---|---|
| ELIXA | 41/3031 (1%) | 35/3032 (1%) | 8·1% | 1.16 (0.74–1.83) |
| LEADER | 87/4668 (2%) | 97/4672 (2%) | 16.5% | 0.89 (0.67–1.19) |
| SUSTAIN-6 | 18/1648 (1%) | 14/1649 (1%) | 3.7% | 1.28 (0.64–2.58) |
| EXSCEL | 246/6456 (4%) | 273/6458 (4%) | 29.6% | 0.88 (0.74–1.05) |
| REWIND | 169/4949 (3%) | 237/4952 (5%) | 25.9% | 0.70 (0.57–0.85) |
| AMPLITUDE-O | 121/2717 (5%) | 76/1359 (6%) | 16.2% | 0.77 (0.57–1.02) |
| **Overall** | **682/22885 (3%)** | **732/21493 (3%)** | **100.0%** | **0.84 (0.73–0.97)** **$P = 0.016$** |

Heterogeneity: $I^2 = 31.4\%$, $P = 0.200$

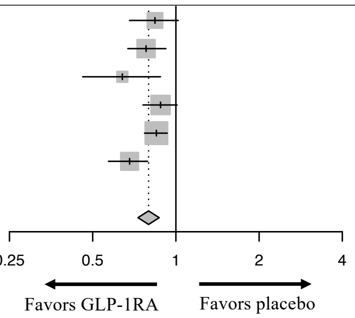

**Supplementary Figure 2**. Subgroup analysis of effects of GLP-1RAs on the primary composite MACE outcome stratified by prior history of established cardiovascular disease.

| Primary Composite MACE Outcome (Established CVD) | Treatment Events/Total (%) | Placebo Events/Total (%) | Weight | Hazard ratio (95% CI) |
|---|---|---|---|---|
| ELIXA | 400/3034 (13%) | 392/3034 (13%) | 15.3 | 1.02 (0.89–1.17) |
| LEADER | 536/3831 (14%) | 692/3736 (19%) | 17.8 | 0.83 (0.74–0.93) |
| SUSTAIN-6 | 98/1353 (5%) | 137/1382 (10%) | 6.6 | 0.72 (0.55–0.93) |
| EXSCEL | 722/5394 (13%) | 786/5388 (15%) | 19.5 | 0.90 (0.82–1.00) |
| HARMONY | 338/4731 (7%) | 428/4732 (9%) | 15.0 | 0.78 (0.68–0.90) |
| PIONEER-6 | 57/1350 (4%) | 68/1345 (5%) | 4.2 | 0.83 (0.58–1.17) |
| REWIND | 280/1560 (18%) | 315/1554 (20%) | 13.1 | 0.87 (0.74–1.02) |
| AMPLITUDE-O | 177/2420 (7%) | 122/1230 (10%) | 8.4 | 0.71 (0.57–0.90) |
| **Overall** | **2608/23673 (11%)** | **2940/22401 (13%)** | **100.0%** | **0.85 (0.79–0.92)** |
| Heterogeneity: $I^2 = 48.0\%$, $P = 0.062$ | | | | ***P* < 0.001** |

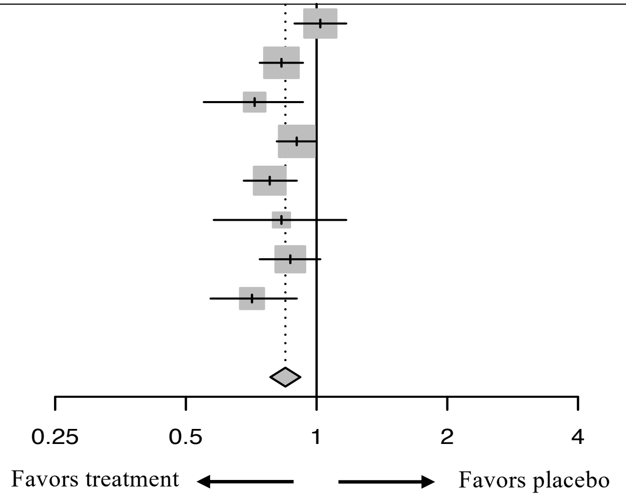


| Primary Composite MACE Outcome (No established CVD) | Treatment Events/Total (%) | Placebo Events/Total (%) | Weight | Hazard ratio (95% CI) |
|---|---|---|---|---|
| LEADER | 72/837 (9%) | 65/905 (7%) | 13.8 | 1.20 (0.86–1.67) |
| SUSTAIN-6 | 10/295 (3%) | 9/267 (3%) | 1.9 | 1.00 (0.41–2.46) |
| EXSCEL | 117/1962 (6%) | 119/2008 (6%) | 23.5 | 0.99 (0.77–1.28) |
| PIONEER-6 | 4/241 (2%) | 8/247 (3%) | 1.0 | 0.51 (0.15–1.68) |
| REWIND | 277/3093 (9%) | 317/3128 (10%) | 58.9 | 0.87 (0.74–1.02) |
| AMPLITUDE-O | 12/297 (4%) | 3/129 (2%) | 0.9 | 1.17 (0.48–6.07) |
| **Overall** | **549/7244 (7%)** | **592/7190 (8%)** | **100.0%** | **0.94 (0.83–1.06)** |
| Heterogeneity: $I^2 = 0.0\%$, p = 0.514 | | | | ***P* = 0.303** |

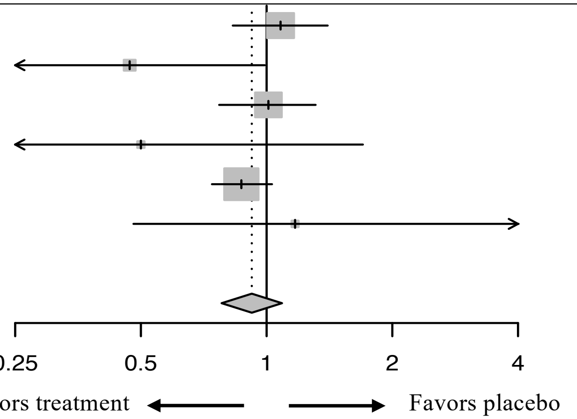


GLP-1RAs = glucagon-like peptide-1 receptor agonists; MACE = major adverse cardiovascular events; CVD = cardiovascular disease.

**Supplementary Figure 3**. Subgroup analysis of effects of GLP-1RAs on the primary composite MACE outcome stratified by structural similarities.

| Primary Composite MACE Outcome (Exendin-based GLP1RAs) | Treatment Events/Total (%) | Placebo Events/Total (%) | Weight | Hazard ratio (95% CI) |
|---|---|---|---|---|
| ELIXA | 400/3034 (13%) | 392/3034 (13%) | 35.3% | 1.02 (0.89–1.17) |
| EXSCEL | 839/7356 (11%) | 905/7396 (12%) | 42.4% | 0.91 (0.83–1.00) |
| AMPLITUDE-O | 189/2717 (7%) | 125/1359 (9%) | 22.4% | 0.73 (0.58–0.92) |
| **Overall** | **1428/13107 (11%)** | **1422/11789 (12%)** | **100.0%** | **0.90 (0.78–1.04)** ***P*** **= 0.163** |

Heterogeneity: $I^2$ = 67.2%, p = 0.047

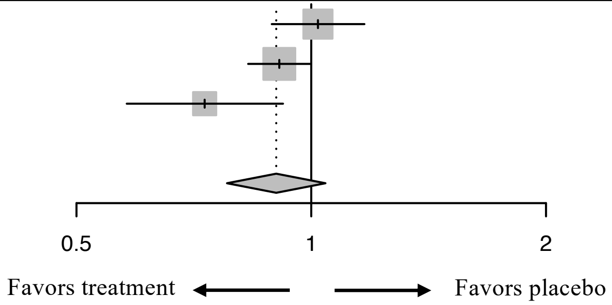


| Primary Composite MACE Outcome (Human GLP-1-based GLP-1RAs) | Treatment Events/Total (%) | Placebo Events/Total (%) | Weight | Hazard ratio (95% CI) |
|---|---|---|---|---|
| LEADER | 608/4668 (13%) | 694/4672 (15%) | 35.2% | 0.87 (0.78–0.97) |
| SUSTAIN-6 | 108/1648 (7%) | 146/1649 (9%) | 6.9% | 0.74 (0.58–0.95) |
| HARMONY | 338/4731 (7%) | 428/4732 (9%) | 21.3% | 0.78 (0.68–0.90) |
| PIONEER 6 | 61/1591 (4%) | 76/1592 (5%) | 3.9% | 0.79 (0.57–1.11) |
| REWIND | 594/4949 (12%) | 663/4952 (13%) | 32.8% | 0.88 (0.79–0.99) |
| **Overall** | **1709/17587 (10%)** | **2007/17597 (11%)** | **100.0%** | **0.84 (0.79–0.90)** ***P*** **< 0.001** |

Heterogeneity: $I^2$ = 0.0%, $P$ = 0.513

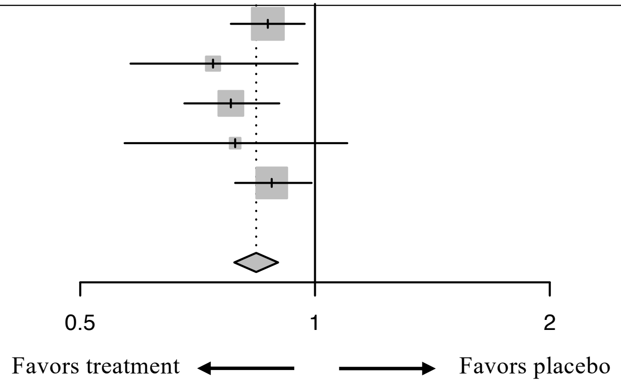


GLP-1RAs = glucagon-like peptide-1 receptor agonists. MACE = major adverse cardiovascular events.

**Supplementary Figure 4.** Univariable meta-regression analysis of normalized reduction in HbA1c (A–C) or body weight (D–F) with the logarithm of hazard ratio (log-HR) for all-cause mortality (A, D), hospitalization due to heart failure (B, E), and renal function outcome (C, F).

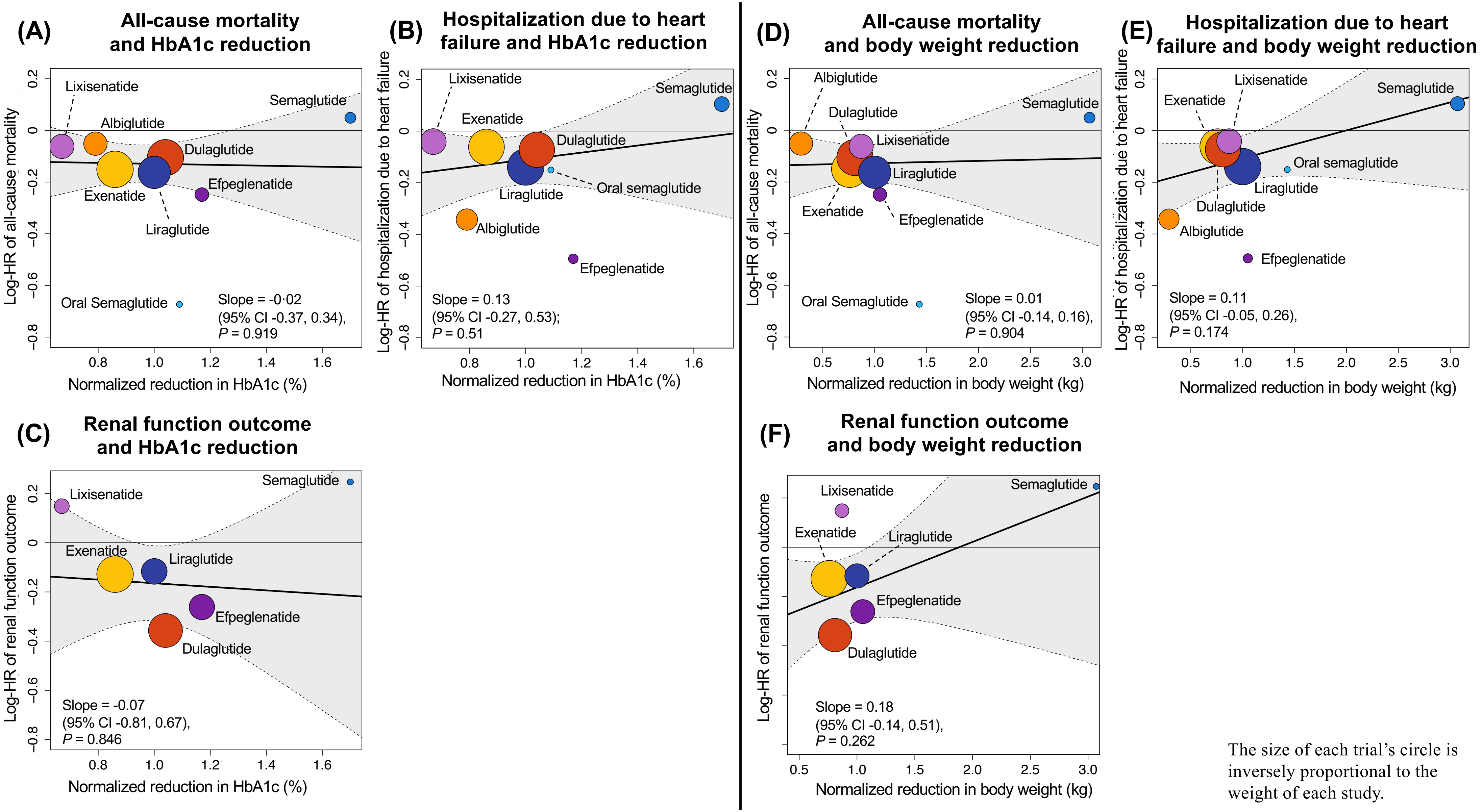


The size of each trial's circle is inversely proportional to the weight of each study.
